# Neuropsychological and neuroanatomical features of patients with behavioral variant Alzheimer’s disease (AD): a comparison to behavioral variant frontotemporal dementia and amnestic AD groups

**DOI:** 10.1101/2022.01.04.21268578

**Authors:** Sophia Dominguez Perez, Jeffrey S. Phillips, Catherine Norise, Nikolas G. Kinney, Prerana Vaddi, Amy Halpin, Katya Rascovsky, David J. Irwin, Corey T. McMillan, Long Xie, Laura E.M. Wisse, Paul A. Yushkevich, Dorina Kallogjeri, Murray Grossman, Katheryn A.Q. Cousins

## Abstract

An understudied non-amnestic variant of Alzheimer’s disease (AD), behavioral variant AD (bvAD) is associated with progressive personality, behavior, or executive dysfunction and frontal atrophy. This study characterizes the neuropsychological and neuroanatomical features associated with bvAD by comparing it to behavioral variant frontotemporal dementia (bvFTD), amnestic AD (aAD), and subjects with normal cognition. Subjects included 16 bvAD, 67 bvFTD, and 18 aAD patients, and 26 healthy controls. Compared to bvFTD, bvAD showed more significant visuospatial impairments (Rey Figure copy and recall), more irritability (Neuropsychological Inventory), and equivalent verbal memory (Philadelphia Verbal Learning Test). Compared to aAD, bvAD indicated more executive dysfunction (F-letter fluency) and better visuospatial performance. Neuroimaging analysis found that bvAD showed cortical thinning relative to bvFTD posteriorly in left temporal-occipital regions; bvFTD had cortical thinning relative to bvAD in left inferior frontal cortex. bvAD had cortical thinning relative to aAD in prefrontal and anterior temporal regions. All patient groups had lower volumes than controls in both anterior and posterior hippocampus. However, bvAD patients had higher average volume than aAD patients in posterior hippocampus and higher volume than bvFTD patients in anterior hippocampus after adjustment for age and intracranial volume. Findings demonstrated that underlying pathology mediates disease presentation in bvAD and bvFTD.

## INTRODUCTION

Alzheimer’s disease (AD) is defined pathologically by the presence of amyloid plaques and neurofibrillary tangles in the brain, which can be detected at autopsy [1,2]. Most common is the amnestic variant of AD (aAD), which presents with impaired episodic memory, executive functioning, naming, and spatial perception, linked to initial atrophy in medial temporal and parietal lobes that ultimately spreads to neocortical association cortices in lateral temporal, parietal and prefrontal regions [3,4]. A rare presentation of AD, however, involves personality and/or executive deficits associated with frontal lobe atrophy, and comparatively spared memory in the beginning of disease course; hereafter we use the acronym bvAD to refer to this variant with behavioral and dysexecutive features [5–9]. Also known as frontal variant AD [10–12], bvAD is difficult to differentiate from the phenotypically similar behavioral variant frontotemporal dementia (bvFTD) [10,13,14]. bvFTD is characterized by progressive deterioration of personality, behavior, social comportment, and executive functioning, related to frontal and anterior temporal lobe atrophy [15]. Although bvFTD and bvAD are clinically similar, the pathologies underlying these syndromes are distinct. While bvFTD is due to frontotemporal lobar degeneration (FTLD) spectrum pathology associated with misfolded tau or TDP-43 [16,17], up to 25% of patients with a clinical diagnosis of bvFTD are found to have AD pathology at autopsy [18,19]. Due to its rarity and clinical overlap with bvFTD, bvAD is difficult to detect and is consequently a poorly defined syndrome that has been understudied. Accurate and non-invasive methods of identifying bvAD can have significant implications for prognosis, caregiver burden, and early intervention [20–24], and might aid development of specialized treatments [16,25]. In this study, we seek to understand how differences in pathology affect neuropsychological and neuroanatomical changes associated with bvAD, bvFTD, and aAD, which could have implications for the distinct clinical needs of each syndrome.

To date, only a few studies have directly compared the clinical characteristics of bvAD and bvFTD, with mixed conclusions [9,26,27]. Differences in findings may be explained by differences in assessments administered, but also by the challenges specific to the clinical population. First, it is a rare condition, resulting in relatively small research sample sizes that may account for discrepant findings. Second, there is no consensus on the definition of bvAD, and previous studies of bvAD have included patients with traumatic brain injury (TBI) [9], with known co-pathologies [27], or who did not meet bvFTD criteria [9,26]. The goal of this study is to build on past work examining the distinct clinical features involved in bvAD associated with AD pathology by comparing it to other conditions. In addition, we aim to investigate differences in anterior and posterior hippocampal volumes in these syndromes. We restricted our bvAD group to patients who met clinical bvFTD criteria [15], who did not have an amnestic deficit at presentation, and who had likely or known AD pathology based on cerebrospinal fluid (CSF) biomarkers or autopsy assessment. To outline a differential profile of bvAD, we tested cognitive, affective, and neuroimaging measures—including neocortical and hippocampal analyses—in bvAD compared to bvFTD, aAD, and healthy controls.

Based on previous findings, we anticipated that bvAD patients would have a partially unique neuropsychological profile, with significant dysexecutive impairment as seen in bvFTD, combined with moderate visuospatial impairment and relatively mild impairment on memory compared to aAD [9,26–28]. For psychiatric and social domains, we expected bvAD to have more signs of anxiety and emotional lability, and bvFTD to have more social symptoms, such as apathy and disinhibition [29]. As seen by previous neuroimaging comparisons, we hypothesized that bvAD patients would show more posterior neocortical atrophy, such as in the temporal and inferior parietal lobes, and medial temporal lobe atrophy, compared to more frontal and anterior temporal atrophy in bvFTD [9,29–31]. Between bvAD and aAD, we expected more frontal atrophy in bvAD [9,26] and more medial temporal atrophy in aAD [30].

## METHODS

### PARTICIPANT SELECTION

The present study is based on a convenience sample of patients and controls who had neuropsychological, magnetic resonance imaging (MRI), and CSF and/or autopsy data in the Integrated Neurodegenerative Disease Database [32] collected between 2005 and 2018. Participants were recruited through the University of Pennsylvania Cognitive Neurology Clinic, from among patients’ caregivers and family, and from the surrounding community. Clinical diagnoses of participants were based on the consensus of a multidisciplinary team using medical history, neurologic examination, and a mental status evaluation. Exclusion criteria included English as a second language and concurrent medical, psychiatric, or neurological (e.g., head trauma, stroke, hydrocephalus) conditions that could alter cognition, behavior, or brain integrity.

### CEREBROSPINAL FLUID

Lumbar punctures were performed as previously described [33]. CSF was collected, aliquoted, and frozen at −80°C within an hour. Samples were assayed via Luminex or ELISA for Aß_42_ and t-tau levels [13]. Seventy subjects had CSF but no autopsy data; of these, 65 had Luminex values, and 5 had ELISA values that were transformed to Luminex equivalents, using an autopsy-confirmed formula [13].

### PARTICIPANT GROUPS

Ninety-six individuals met clinical diagnostic criteria for bvFTD [15], and were divided into bvAD and bvFTD groups. From these subjects, we excluded six who showed mixed AD and FTLD pathology at autopsy, two with secondary or tertiary Lewy body pathology at autopsy, and five with a concurrent diagnosis of amyotrophic lateral sclerosis or corticobasal degeneration. Of the remaining 83 subjects, 21 met definite bvFTD criteria, 38 met probable criteria, and 24 met possible criteria for bvFTD.

Those with AD or FTLD pathology were classified as bvAD or bvFTD, respectively, based on results from autopsy (performed as previously described [19]; n=13) if available, or based on CSF t-tau/Aβ_42_ ratio (n=70). Using a pre-defined CSF t-tau/Aβ_42_ cut-off [13], individuals with a ratio > 0.34 were classified as bvAD, and those with a ratio < 0.34 were classified as bvFTD. Our final cohort included 16 bvAD and 67 bvFTD participants. Three bvAD and seven bvFTD patients included in the final groups had a secondary clinical diagnosis of semantic variant primary progressive aphasia (svPPA). All patients with aAD met the pre-defined CSF threshold (t-tau/Aβ_42_ ratio > 0.34) [13], confirming likely AD pathology.

In addition, bvAD patients were compared to patients with typical aAD (n=18) and elderly healthy controls (n=26) that had no self-reported known neurological or psychiatric disorders. Mini-Mental Status Examination (MMSE) scores, used a proxy for disease severity, were above 15 for aAD and 27 for healthy control participants.

### NEUROPSYCHOLOGICAL ASSESSMENTS

Subsets of neuropsychological data available for each patient were used to profile multiple cognitive and behavioral domains associated with bvFTD: attention and executive function, verbal memory, visuospatial skills and memory, and psychiatric and social symptoms. The first cognitive domain assessed attention and executive function. Forward digit span, requiring repetition of a sequence of digits, measured immediate attention [34]. Executive function was measured with backward digit span, requiring repetition of digit spans in the reverse order (working memory), and number of F-words named in one minute (fluency and inhibition) [35]. Second, the Philadelphia Verbal Learning Test (PVLT) [36] measured verbal memory and recognition by testing subjects’ ability to learn and remember a list of nine words on multiple trials. The long delay recall score represents the number of words retained and correctly recalled after a 10-15-minute delay. The recognition score was the number of correct words recognized from a list of 9 target words and 27 lures. Third, patients were asked to copy and recall the Rey Figure [37,38], a task involving visuoperceptual, visuoconstructional, and visual memory functions [39,40]. Finally, we used the Neuropsychiatric Inventory (NPI), an informant-based interview, to evaluate psychiatric and social symptoms [41,42]. NPI data in the current study combined scores from the NPI and a brief questionnaire form of the NPI (NPI-Q) to measure the presence of 12 symptoms: delusions, hallucinations, agitation/aggression, depression/dysphoria, anxiety, elation/euphoria, apathy/indifference, disinhibition, irritability/emotional lability, motor disturbance (gauging repetitive movements), nighttime behaviors, and appetite/eating. We compared differences in the presence of NPI symptoms between groups, as well as the total NPI score. While no NPI data were available for controls, the presence of a symptom indicates a deviation from the norm.

### STATISTICAL ANALYSES FOR NEUROPSYCHOLOGICAL ASSESSMENTS

Kruskal-Wallis tests performed non-parametric comparisons across groups for age at onset (age at earliest reported symptom), age at test, disease duration (interval from onset to test), and MMSE.

Neuropsychological data were not normally distributed according to Shapiro-Wilks tests (all p<0.05). We performed non-parametric comparisons by rank-transforming continuous neuropsychological data among the four groups [43]. One-way analysis of covariance (ANCOVA) tested differences across groups for each rank-transformed cognitive measure. Logistic regressions evaluated the presence or absence of each behavioral disturbance (NPI) in bvAD compared to aAD and bvFTD. To account for effects of age and disease severity, ANCOVAs and logistic regressions included age at test and MMSE as covariates. Statistical models were also tested without covariates to ensure statistical robustness; these reduced models yielded no change to statistical significance except for the Rey Copy task, as noted in the results. We report results from models that included covariates. Because we are evaluating a rare population, we use the liberal detection threshold of α = 0.05 two-tailed for all tests.

### NEUROIMAGING METHODS

A large subset of bvAD and bvFTD patients (n=80) had MRI scans within a year of neuropsychological data and/or CSF. Participants completed an axial T1-weighted magnetization-prepared rapid gradient echo (MPRAGE) scan acquired at the Hospital of the University of Pennsylvania on a Siemens 3.0 Tesla scanner with an eight-channel coil using the following parameters: repetition time (TR)=1,620 ms; echo time (TE)=3 ms; flip angle=15°; matrix size=192 × 256; in-plane resolution=0.9766 mm x 0.9766 mm; and slice thickness=1 mm. Images were visually inspected, and scans with head motion or other artifacts were excluded. For analysis of whole-brain cortical thickness, image processing was performed with the Advanced Normalization Tools (ANTs) [44,45] software package. All images were intensity-normalized [46] and spatially normalized to a custom template derived from healthy controls scans from the Open Access Series of Imaging Studies (OASIS) dataset [47] using a symmetric diffeomorphic algorithm [48,49]. Priors from the template image were used to segment each brain into cortical grey matter, subcortical grey matter, deep white matter, CSF, brainstem, and cerebellum; cortical thickness was then estimated on the basis of this tissue segmentation [45]. Voxelwise group analyses were performed by warping cortical thickness images to the template, spatially smoothing them with a Gaussian kernel of 4 mm full-width half-maximum, and downsampling them to 2 mm isotropic voxels. Because hippocampal grey matter is not well-segmented using standard pipelines for cortical thickness estimation [50–52], we used a segmentation algorithm [53] based on the Automated Segmentation of Hippocampal Subfields (ASHS) method [54] to estimate hippocampal volume from participants’ T1-weighted MRIs. This algorithm uses priors defined from expert manual segmentations to divide the medial temporal lobe in each hemisphere into the anterior and posterior hippocampus, adjacent cortical areas, the collateral and occipitotemporal sulci, and meninges. Two raters (JSP and NGK) visually confirmed segmentation quality by overlaying the ASHS and ANTs segmentations on each participant’s T1-weighted image; mislabeling of ventricular CSF as hippocampus was found in ten images and corrected by manually editing hippocampal segmentations.

Voxelwise differences in cortical thickness were evaluated by two-sample unpaired t-tests between groups, covarying for age, sex, and MMSE. Statistical significance was assessed non-parametrically using FSL’s randomise for each contrast, based on 5,000 permutations of the input data [55]. Cortical thickness in the bvAD, bvFTD, and aAD groups was assessed relative to controls at a significance level of p<0.05, after application of FSL’s threshold-free cluster enhancement (TFCE) algorithm [56] and family-wise error correction for multiple comparisons. In a direct contrast of the bvAD and bvFTD groups, no results were observed after multiple comparisons correction; exploratory results are presented at an uncorrected threshold of p<0.001 without TFCE. All voxelwise results are presented with a minimum cluster volume of 25 voxels (200 µl). Anterior and posterior hippocampal volumes were contrasted between groups using linear mixed effects models for each region covarying for age, sex, MMSE; additionally, we covaried for intracranial volume to account for interindividual differences in head size. A random intercept per participant was included to account for within-participant correlation of hippocampal volume estimates. We used model coefficients to adjust for significant covariate effects, and two-sample t-tests with Welch’s adjustment for unequal variance and false discovery rate (FDR) correction were used to perform pairwise group contrasts of adjusted volumes; a criterion of p<0.05 was used to determine statistical significance.

### STANDARD PROTOCOL APPROVALS, REGISTRATIONS, AND PATIENT CONSENTS

All procedures, including CSF collection, MRI, neuropsychological testing, and required informed consent were performed in accordance with the Declaration of Helsinki and the rules of the Institutional Review Board at the University of Pennsylvania.

## RESULTS

### DEMOGRAPHIC COMPARISONS

Clinical and demographic information for all groups is summarized and compared in Table 1. Across bvAD, bvFTD, aAD, and healthy control groups, non-parametric Kruskal-Wallis tests compared continuous variables and a chi-square test compared sex and race. Groups were not significantly different on education or sex (both p>0.1), but had significantly different ages at test (H(3)=8.09, p=0.04), MMSE scores (H(3)=47.9, p<0.001), and race (*χ*^2^(9)=22.75, p=0.007). Patients (bvFTD, bvAD, and aAD) were not significantly different for disease duration (p>0.1), but had significantly different ages of onset (H(2)=6.62, p=0.04). Post-hoc test indicated that bvAD patients had significantly older age at test (W=743.5, p=0.02) and significantly lower MMSE scores (W=349, p=0.03) than bvFTD patients. Therefore, comparisons of neuropsychological assessments covary for age at test and MMSE.

**Table 1.** Clinical and demographic summary, showing median (M), interquartile interval (IQI), and sample size (n) for age at MRI, age at onset, education, disease duration, and MMSE. Sex indicates the number of males (m) and females (f). Race/Ethnicity indicates number of White (W), Black (B), Asian (A), and multi-racial (M) participants; superscripts indicate number of Latinx (L) participants within each racial group. Comparison column summarizes results of Kruskal-Wallis and chi-square tests comparing all groups, with asterisks indicating group differences: *p<0.05, **p<0.01, ***p<0.001. Asterisks by group medians indicate significant difference from bvAD for age at test, age at onset, education, disease duration, and MMSE.

| Demographics |  | bvAD | bvFTD | aAD | Healthy Control | Comparisons |  |
| --- | --- | --- | --- | --- | --- | --- | --- |
| | | | | | | $\chi$ | $p$ |
| **Age at Test**<br>(years) | M | 68 | 62* | 68.5 | 61 | 8.09 | 0.04* |
|  | IQI | 59.75-72 | 57-67 | 57.5-73 | 57.75-68.75 |  |  |
|  | n | 16 | 67 | 18 | 26 |  |  |
| **Age at Onset**<br>(years) | M | 63.5 | 59 | 63.5 | - | 6.62 | 0.04* |
|  | IQI | 55.5-69 | 53.5-65 | 56-70 | - |  |  |
|  | n | 16 | 67 | 18 | - |  |  |
| **Education**<br>(years) | M | 17 | 16 | 17 | 16 | 2.24 | 0.53 |
|  | IQI | 15-18 | 12-18 | 14.5-18.75 | 13.25-18 |  |  |
|  | n | 16 | 67 | 18 | 26 |  |  |
| **Disease Duration**<br>(years) | M | 3 | 3 | 2.5 | - | 1.32 | 0.52 |
|  | IQI | 2-5 | 1-4.5 | 1-3.75 | - |  |  |
|  | n | 16 | 67 | 18 | - |  |  |
| **MMSE Score**<br>(points out of 30) | M | 23 | 26* | 23 | 30*** | 47.9 | 2e-10<br>*** |
|  | IQI | 19.75-26 | 23.5-28 | 18.75-25 | 28-30 |  |  |
|  | n | 16 | 67 | 18 | 23 |  |  |
| **Sex** | m | 12 | 42 | 9 | 11 | 5.61 | 0.13 |
|  | f | 4 | 25 | 9 | 15 |  |  |
| **Race/Ethnicity** | W | 15 | 62 | 16<sup>L=1</sup> | 18 | 22.75 | 0.007<br>** |
|  | B | 1 | 2 | 0 | 7 |  |  |
|  | A | 0 | 3 | 1 | 0 |  |  |
|  | M | 0 | 0 | 1<sup>L=1</sup> | 1 |  |  |

### NEUROPSYCHOLOGICAL TESTING

Results from ANCOVAs between control and patient groups are outlined in Table 2 and summarized in Figure 1. We found a main effect of group for all cognitive measures within each domain (all p<0.05). Planned ANCOVA comparisons directly compared bvAD to bvFTD, aAD, and healthy controls. When compared to controls, bvAD patients performed significantly worse on all tasks (all p<0.05). Comparing bvAD and bvFTD, we found no statistical differences (all p>0.1) for attention and executive function tasks (Digit Forward Span, Digit Backward Span, F-words) or verbal memory (PVLT delayed recall or recognition). On visuospatial skills and visual memory, bvAD patients performed significantly worse than bvFTD patients, with lower scores on both Rey Figure copy [F(1,59)=5.52, p=0.02] and recall [F(1,57)=5.73, p=0.02]. When compared to aAD, bvAD patients performed worse on one executive function task [F-words: F(1,22)=6.63, p=0.02] and performed better on visuospatial tasks [Rey Figure copy: F(1,21)=4.77, p=0.04; Rey Figure recall: F(1,20)=9.11, p=0.007]. No differences were found between bvAD and aAD on verbal memory (p>0.1).

**Table 2.** Median (M), interquartile interval (IQI), and n for all groups on three cognitive domains. Comparison column summarizes ANCOVA, with asterisks indicating group differences: *p<0.05, **p<0.01, ***p<0.001. Asterisks by group medians indicate significant difference from bvAD

| Cognitive Assessment |  | bvAD | bvFTD | aAD | Healthy Control | Comparisons |  |
| --- | --- | --- | --- | --- | --- | --- | --- |
|  |  |  |  |  |  | F | p |
| Attention and Executive Function |  |  |  |  |  |  |  |
| **Digit Forward Span**<br>(number of digits in longest span correct) | M | 6 | 6 | 6 | 7*** | 5.95 | 0.001 ** |
|  | IQI | 5-6 | 6-7 | 5-7 | 7-8 |  |  |
|  | n | 13 | 43 | 12 | 15 |  |  |
| **Digit Backward Span**<br>(number of digits in longest span correct) | M | 4 | 4 | 3 | 5** | 3.83 | 0.01 * |
|  | IQI | 3-4 | 3-5 | 3-4 | 4.5-6.5 |  |  |
|  | n | 13 | 42 | 12 | 15 |  |  |
| **F-words**<br>(number of words) | M | 8 | 7 | 12* | 16.5** | 10.21 | 1e-05 *** |
|  | IQI | 4-11 | 4-11 | 9-13.5 | 12.5-18 |  |  |
|  | n | 14 | 42 | 12 | 14 |  |  |
| Verbal Memory |  |  |  |  |  |  |  |
| **PVLT Long Delay Recall**<br>(number of words) | M | 3 | 4 | 1 | 7*** | 12.32 | 1e-06 *** |
|  | IQI | 0-4.5 | 1-6 | 0-2 | 7-7.5 |  |  |
|  | n | 15 | 47 | 11 | 11 |  |  |
| **PVLT Recognition**<br>(number of words) | M | 8 | 8 | 7 | 9* | 3.09 | 0.03 * |
|  | IQI | 6.5-9 | 7-9 | 6-8 | 8.5-9 |  |  |
|  | n | 15 | 48 | 11 | 11 |  |  |
| Visuospatial Skills and Memory |  |  |  |  |  |  |  |
| **Rey Copy**<br>(points out of 36) | M | 25 | 32* | 12.5* | 36** | 11.9 | 2e-06 *** |
|  | IQI | 11.12-33.25 | 27-34 | 5.5-25 | 32.25-36 |  |  |
|  | n | 14 | 49 | 11 | 10 |  |  |
| **Rey Recall**<br>(points out of 36) | M | 6.75 | 14.5* | 1** | 17.75*** | 13.67 | 3e-07 *** |
|  | IQI | 2.63-11.88 | 5.75-19.5 | 0-1 | 15.62-23.88 |  |  |
|  | n | 14 | 47 | 10 | 10 |  |  |

**Figure 1.**
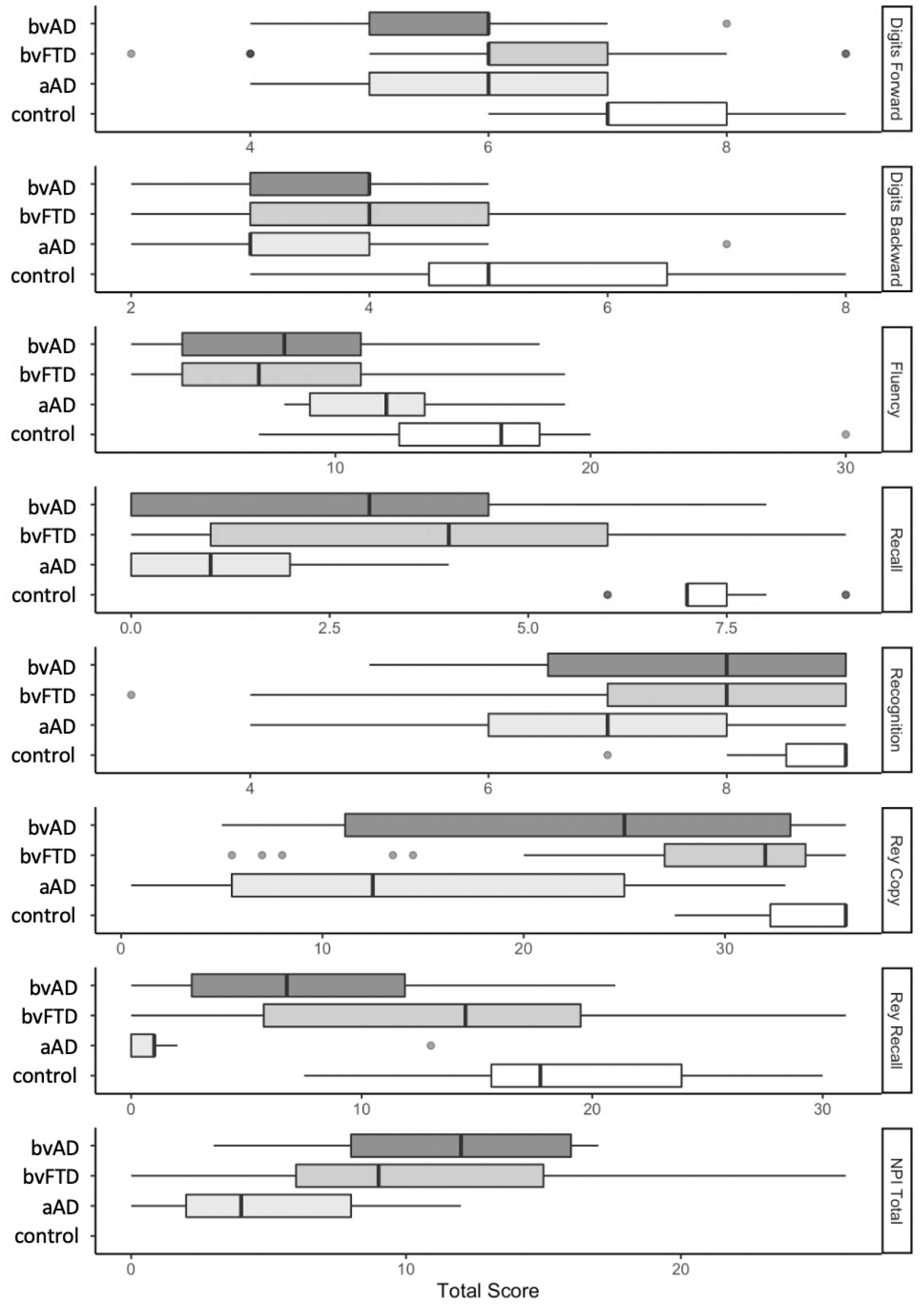
Boxplots comparing bvAD, bvFTD, aAD, and controls on all continuous neuropsychological measures. Measures of attention and executive function include Digits Forward, Digits Backward, and Fluency (F-words). Verbal memory assessments include Recall and Recognition. Visual skills and memory measures include Rey Copy and Recall. Psychiatric and social symptoms are compared with the NPI total score. No data was collected for controls on the NPI.

Logistic regression tested differences in the presence of behavioral disturbances in bvAD compared to bvFTD and aAD. On behavioral NPI measures, caregivers rated bvAD patients as being significantly more likely to exhibit agitated or aggressive behaviors than bvFTD patients (p=0.03), while no difference was found for delusions, hallucinations, depression, anxiety, euphoria, apathy, disinhibition, irritability, motor disturbance, nighttime behaviors, appetite, or NPI total (all p>0.1; Table 3). When compared to aAD, bvAD patients were rated as having significantly higher rates of disinhibition (p=0.006) and motor disturbances (p=0.01), and marginally higher rates of agitation/aggression (p=0.06). All other NPI measures were not significantly different (all p>0.1).

**Table 3.**
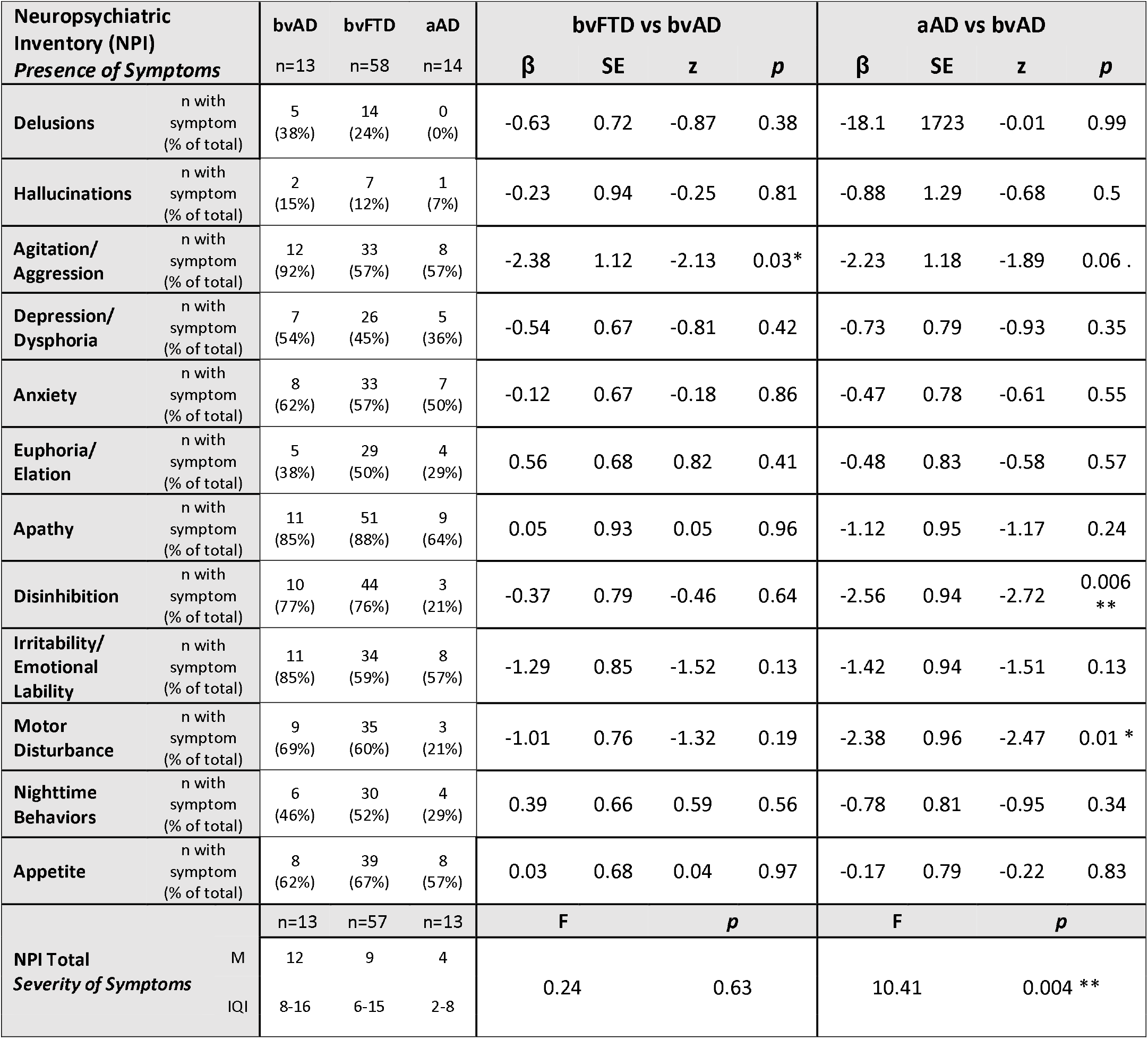
Results of logistic regression comparing each NPI symptom across patient groups (bvAD, bvFTD, aAD). Beta estimate (β), standard error (SE), z-value (z), and p-value (*p*) are relative to bvAD performance. Period and asterisks indicate difference from bvAD:. *p*=0.06, \**p*<0.05, \*\**p*<0.01.

### NEUROIMAGING

Voxelwise contrasts relative to elderly controls revealed extensive cortical thinning in bvAD, bvFTD, and aAD (Table 4, Figure 2). After statistical adjustment for age, sex, and MMSE, patient groups displayed cortical thinning throughout the brain at a significance level of p<0.05 (family-wise error corrected) with relatively spared primary visual and sensorimotor cortices.

**Table 4.** Subpeak voxel locations where patients exhibited greater atrophy (i.e., lower cortical thickness) than healthy controls. Results are based on a statistical threshold of p<0.05, after application of FSL’s threshold-free cluster enhancement algorithm and family-wise error correction for multiple comparisons. Top ten most significant subpeaks in both hemispheres are reported in MNI coordinates.

| Hemisphere | Region | T.statistic | X | Y | Z |
| --- | --- | --- | --- | --- | --- |
| **bvAD vs Healthy Control** |  |  |  |  |  |
| Left | Anterior cingulate gyrus | 5.415 | -2 | 47 | 22 |
|  | Gyrus rectus | 5.408 | -8 | 22 | -22 |
|  | Middle frontal gyrus | 5.241 | -41 | 45 | 12 |
|  | Middle temporal gyrus | 5.213 | -62 | -53 | 1 |
|  | Superior frontal gyrus | 5.19 | -22 | 59 | 7 |
|  | Amygdala | 4.8 | -29 | -7 | -17 |
|  | Anterior insula | 4.756 | -40 | 17 | -3 |
|  | Superior frontal gyrus | 4.67 | -11 | 34 | 50 |
|  | Middle frontal gyrus | 4.486 | -41 | 16 | 43 |
|  | Middle temporal gyrus | 4.453 | -51 | -5 | -29 |
| Right | Superior temporal gyrus | 5.999 | 57 | -37 | 11 |
|  | R anterior insula | 5.915 | 41 | -3 | 11 |
|  | R middle frontal gyrus | 5.836 | 43 | 47 | 10 |
|  | R superior frontal gyrus | 5.433 | 25 | 37 | 40 |
|  | R anterior insula | 5.425 | 42 | 19 | -2 |
|  | Angular gyrus | 5.353 | 44 | -57 | 28 |
|  | Cerebral White Matter | 5.338 | 25 | -6 | -11 |
|  | R middle temporal gyrus | 5.126 | 66 | -19 | -18 |
|  | R superior frontal gyrus | 5.112 | 23 | 52 | 26 |
|  | Posterior orbital gyrus | 5.074 | 34 | 36 | -10 |
| **bvFTD vs Healthy Control** |  |  |  |  |  |
| Left | Anterior insula | 8.722 | -35 | 23 | -3 |
|  | Superior frontal gyrus medial | 8.554 | -8 | 49 | 22 |
|  | Middle frontal gyrus | 8.129 | -34 | 45 | 28 |
|  | Middle frontal gyrus | 7.927 | -41 | 46 | 9 |
|  | Amygdala | 7.713 | -27 | -8 | -12 |
|  | Anterior orbital gyrus | 7.329 | -19 | 63 | -7 |
|  | Middle frontal gyrus | 7.121 | -36 | 21 | 47 |
|  | Entorhinal area | 6.953 | -26 | -3 | -34 |
|  | Superior frontal gyrus | 6.867 | -8 | 49 | 45 |
|  | Temporal pole | 6.773 | -36 | 22 | -36 |
| Right | Anterior insula | 9.353 | 42 | 19 | -2 |
|  | Middle frontal gyrus | 9.09 | 43 | 47 | 10 |
|  | Posterior insula | 9.088 | 44 | -2 | -2 |
|  | Posterior orbital gyrus | 8.32 | 27 | 30 | -20 |
|  | Amygdala | 7.985 | 25 | -6 | -11 |
|  | Middle temporal gyrus | 7.791 | 46 | 0 | -31 |
|  | Middle temporal gyrus | 7.777 | 64 | -28 | -1 |
|  | Middle frontal gyrus | 7.774 | 44 | 37 | 28 |
|  | Superior frontal gyrus | 7.476 | 25 | 60 | 23 |
|  | Temporal pole | 7.375 | 23 | 17 | -38 |
| **aAD vs Healthy Control** |  |  |  |  |  |
| Left | Amygdala | 6.286 | 25 | 10 | -10 |
|  | Middle temporal gyrus | 5.621 | 66 | 46 | 3 |
|  | Middle frontal gyrus | 4.998 | 35 | -47 | 23 |
|  | Precentral gyrus | 4.873 | 45 | -4 | 28 |
|  | Superior occipital gyrus | 4.727 | 24 | 91 | 16 |
|  | Middle frontal gyrus | 4.713 | 36 | -21 | 47 |
|  | Posterior insula | 4.468 | 29 | 27 | 16 |
|  | Angular gyrus | 4.4 | 38 | 62 | 55 |
|  | Supramarginal gyrus | 4.364 | 50 | 55 | 23 |
|  | Superior parietal lobule | 4.276 | 30 | 41 | 48 |
| Right | Middle frontal gyrus | 4.743 | -43 | -47 | 10 |
|  | Amygdala | 4.592 | -27 | 8 | -10 |
|  | Supramarginal gyrus | 4.566 | -48 | 42 | 52 |
|  | Postcentral gyrus | 4.524 | -66 | 17 | 22 |
|  | Angular gyrus | 4.512 | -54 | 55 | 26 |
|  | Posterior insula | 4.425 | -44 | 4 | 1 |
|  | Inferior occipital gyrus | 4.277 | -33 | 86 | -4 |
|  | Posterior cingulate gyrus | 4.255 | -1 | 42 | 35 |
|  | Superior parietal lobule | 4.184 | -24 | 63 | 45 |
|  | Superior temporal gyrus | 4.169 | -64 | 36 | 10 |

**Figure 2.**
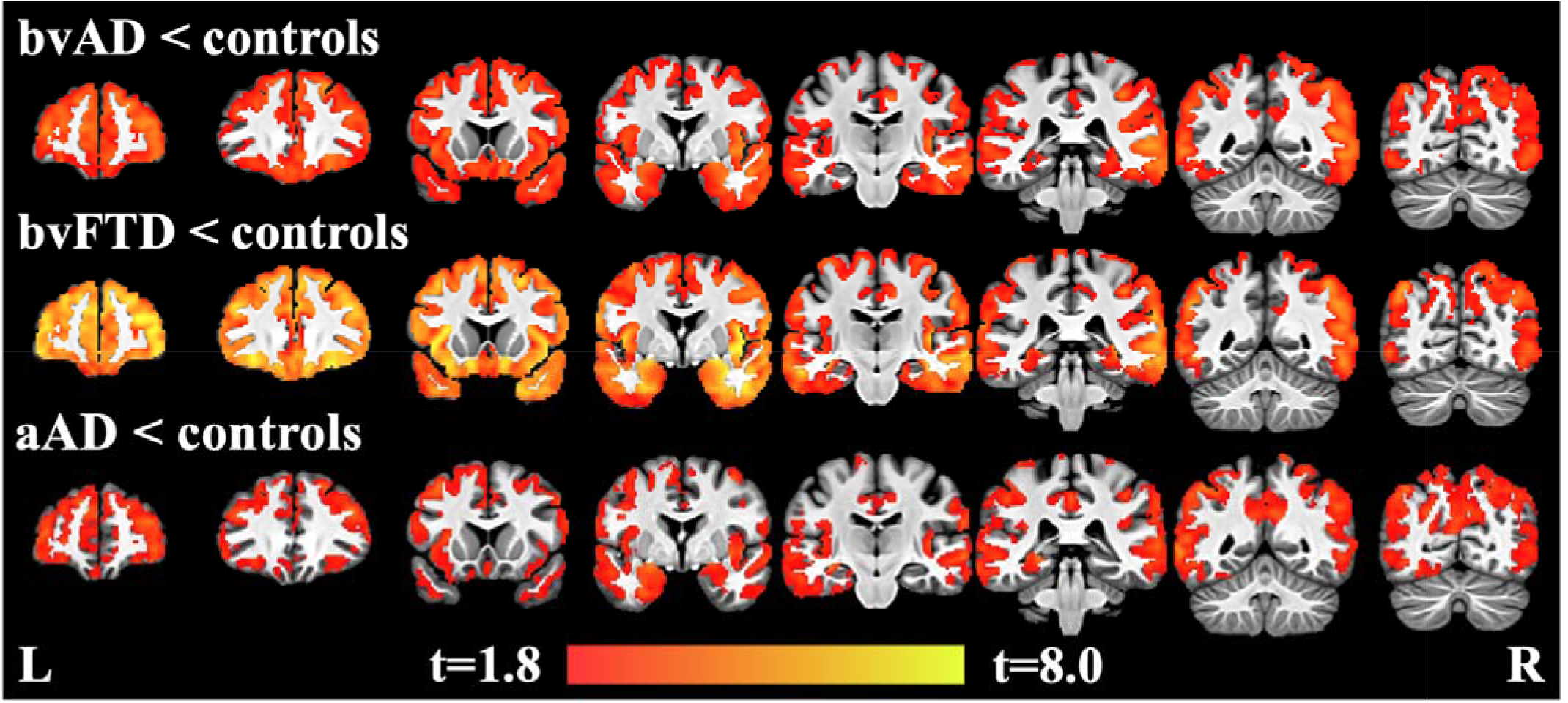
Regions of decreased cortical thickness in bvAD, bvFTD, and aAD compared to controls, covarying for age, sex, and MMSE, and using family-wise error corrected significance of p<0.05 and a cluster threshold of 25 voxels (200 µl). Heatmap indicates t-values.

In a direct contrast of bvAD and bvFTD patients, no significant differences in cortical thickness were observed after correction for multiple comparisons. However, at p<0.001 (uncorrected) with a minimum cluster volume of 200 µl, exploratory analyses showed that the bvAD group had cortical thinning relative to the bvFTD group in right superior temporal cortex, left fusiform gyrus, and left central operculum (Figure 3; Table 5). Conversely, the bvFTD group had cortical thinning relative to the bvAD group in left inferior frontal gyrus (pars orbitalis).

**Figure 3.**
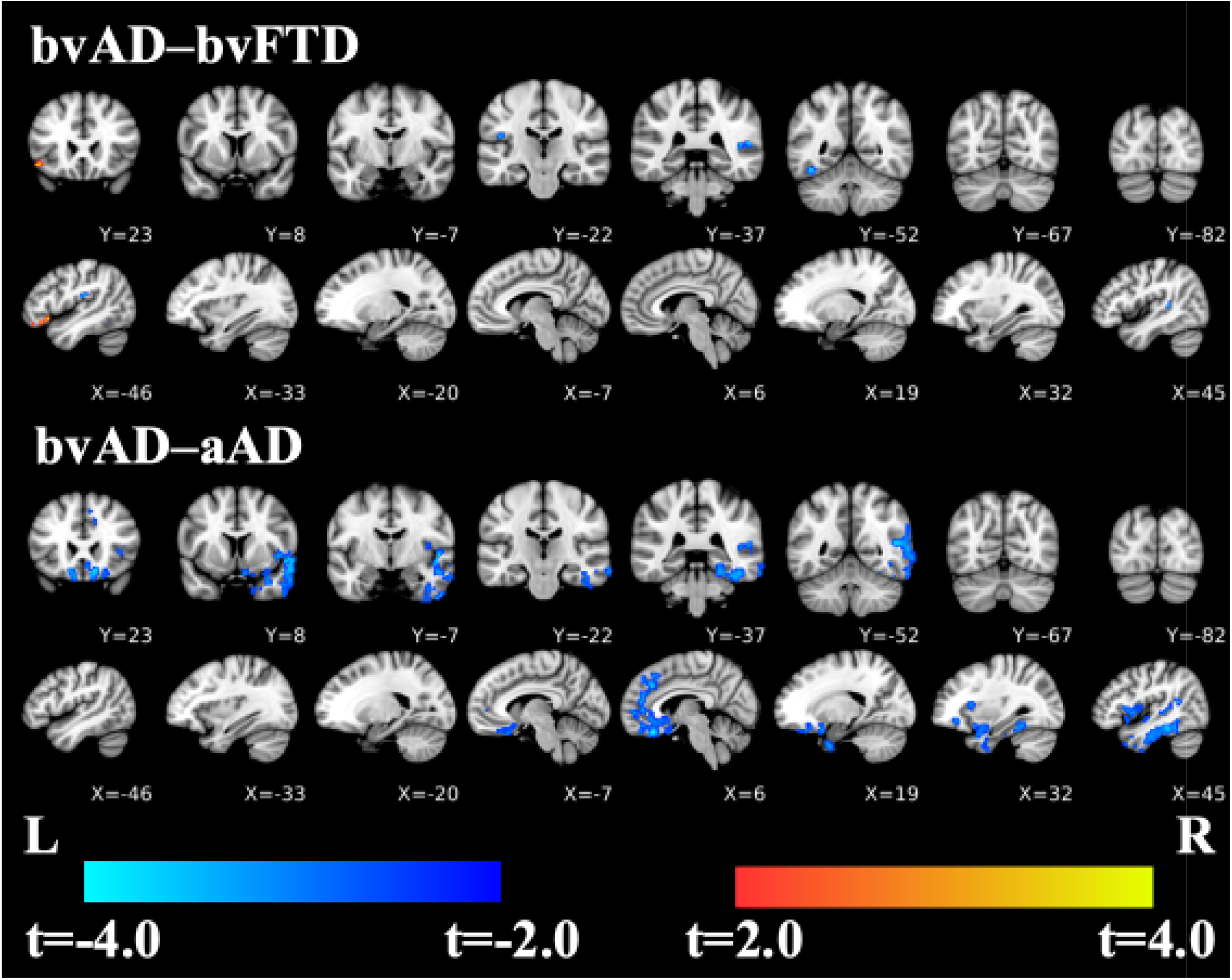
Contrasts of bvAD relative to bvFTD and aAD, covarying for age, sex, and MMSE, using TFCE with an uncorrected significance of p<0.001 and a cluster threshold of 25 voxels (200 µl). Cool colors indicate where bvAD has cortical thinning relative to the bvFTD and aAD groups; warm colors indicate where bvFTD group has cortical thinning compared to the bvAD group (left inferior frontal gyrus, first coronal and sagittal slices). There are no such regions where bvAD>aAD.

**Table 5.**
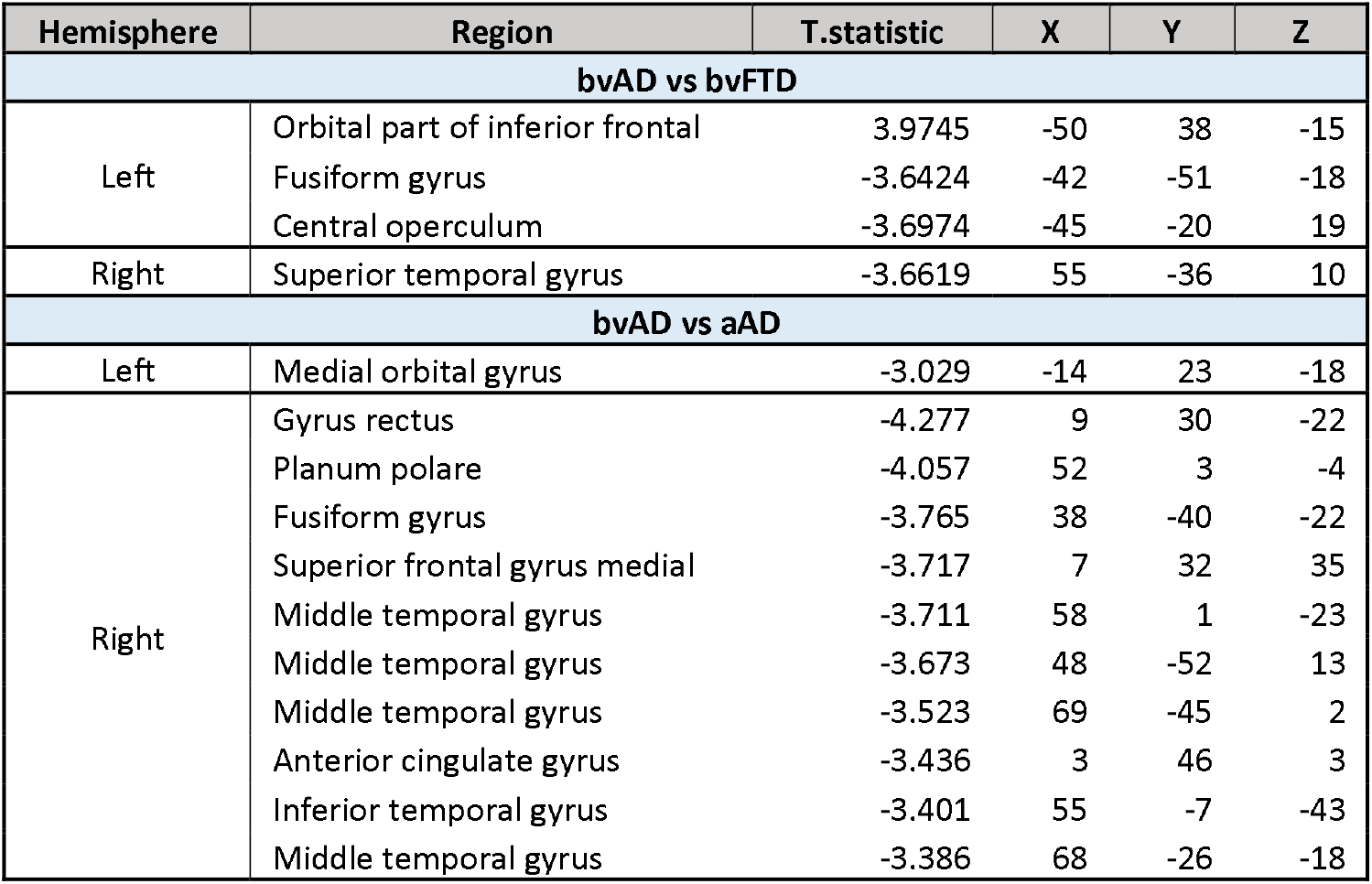
bvAD compared to bvFTD (using peak clusters) and aAD (using subpeak clusters). Results are based on a voxelwise statistical threshold of p<0.001, uncorrected for multiple comparisons. Negative T-statistic values indicate regions where bvAD has cortical thinning relative to a comparison group.

| Hemisphere | Region | T.statistic | X | Y | Z |
| --- | --- | --- | --- | --- | --- |
| **bvAD vs bvFTD** |  |  |  |  |  |
| Left | Orbital part of inferior frontal | 3.9745 | -50 | 38 | -15 |
|  | Fusiform gyrus | -3.6424 | -42 | -51 | -18 |
|  | Central operculum | -3.6974 | -45 | -20 | 19 |
| Right | Superior temporal gyrus | -3.6619 | 55 | -36 | 10 |
| **bvAD vs aAD** |  |  |  |  |  |
| Left | Medial orbital gyrus | -3.029 | -14 | 23 | -18 |
| Right | Gyrus rectus | -4.277 | 9 | 30 | -22 |
|  | Planum polare | -4.057 | 52 | 3 | -4 |
|  | Fusiform gyrus | -3.765 | 38 | -40 | -22 |
|  | Superior frontal gyrus medial | -3.717 | 7 | 32 | 35 |
|  | Middle temporal gyrus | -3.711 | 58 | 1 | -23 |
|  | Middle temporal gyrus | -3.673 | 48 | -52 | 13 |
|  | Middle temporal gyrus | -3.523 | 69 | -45 | 2 |
|  | Anterior cingulate gyrus | -3.436 | 3 | 46 | 3 |
|  | Inferior temporal gyrus | -3.401 | 55 | -7 | -43 |
|  | Middle temporal gyrus | -3.386 | 68 | -26 | -18 |

The bvAD group had cortical thinning relative to the aAD group in right temporal (planum polare; inferior and middle temporal gyri; and fusiform gyrus) and right prefrontal (gyrus rectus, anterior cingulate gyrus, and medial superior frontal gyrus) cortex as well as the left medial orbital frontal gyrus (Figure 3; Table 5). aAD patients did not have smaller cortical thickness than bvAD in any brain regions.

Group differences in anterior and posterior hippocampal volumes are visualized in Figure 4 and Table 6. ANOVAs on linear mixed effects models indicated main effects of group in both anterior [F(3, 107)=6.03, p=0.0008] and posterior hippocampus [F(3, 107)=9.95, p<0.0001]. Intracranial volume was also associated with volume in both regions [anterior: F(1, 107)=32.06, p<0.0001; posterior: F(1,107)=29.59, p<0.0001]; age effects were significant in anterior hippocampus [F(1,107)=4.86, p=0.03] and marginal in posterior hippocampus [F(1,107)=3.35, p=0.07]. Associations with sex and MMSE score were non-significant in both regions (all F<0.8, p>0.38). Hippocampal volumes were thus adjusted for age and intracranial volume.

**Figure 4.**
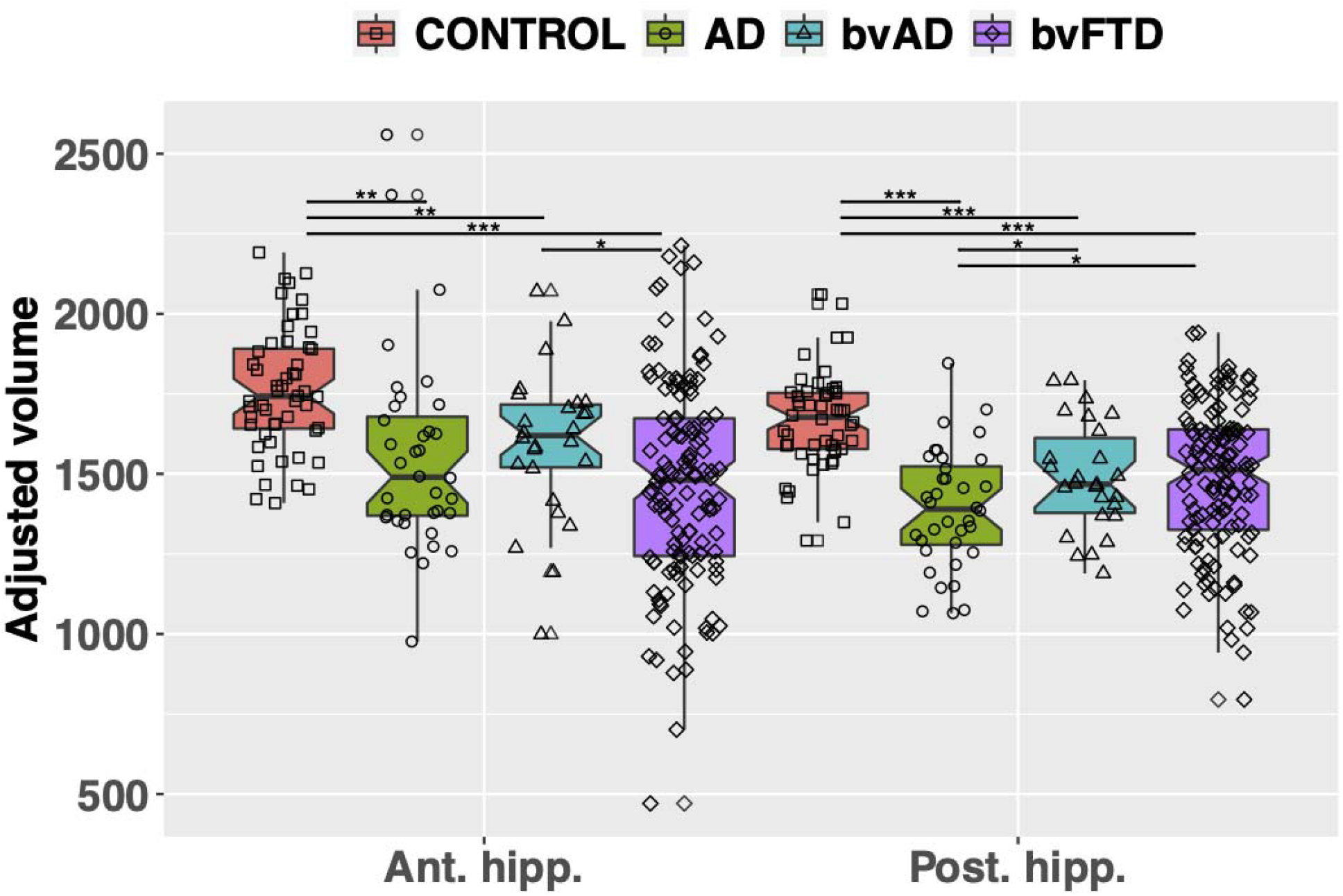
Group differences in anterior and posterior hippocampal volume, adjusted for age and intracranial volume. Volumes are in microliters, normalized by the number of slices imaged in each participant. Significant between-group differences are marked by lines with asterisks: * = p<0.05; ** = p<0.01; *** = p<0.001.

**Table 6.**
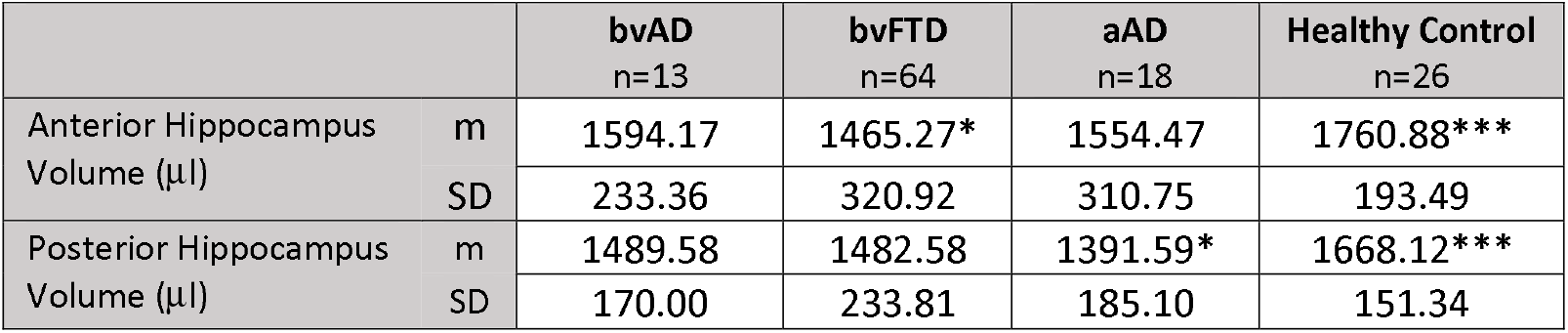
Mean (m), standard deviation (SD), and n for all groups on anterior and posterior hippocampal volume, adjusted for effects of age and intracranial volume. Asterisks by group means indicate significant difference from bvAD based on post-hoc two-sample t-tests with FDR correction: *p<0.05, **p<0.01, **p<0.001. Additionally, both bvFTD and aAD had significantly smaller volumes compared to controls in both the anterior and posterior hippocampus.

In post-hoc tests, adjusted volumes for the bvAD group were significantly lower than controls’ in both anterior [t(42.7)=-3.14, p=0.006] and posterior hippocampus [t(45.3)=-4.53, p=0.0001). In the bvFTD group, volumes were lower than controls’ in both anterior [t(152.3)=-7.57, p<0.0001] and posterior hippocampus [t(143.6)=-6.30, p<0.0001]. While bvAD and bvFTD had comparable volumes in the posterior hippocampus [t(46.5)=0.18, p=0.86], bvFTD had significantly smaller anterior hippocampal volumes than bvAD [t(46.5) = −2.39, p=0.031). Furthermore, aAD had significantly smaller volumes compared to controls in both the anterior [t(53.7)=-3.54, p=0.002] and posterior hippocampus [t(65.3)=-7.41, p<0.0001]. bvAD patients had significantly higher volumes than aAD patients in posterior hippocampus [t(56.5)=2.16, p=0.047] but not in anterior hippocampus [t(59.9)=0.57, p=0.62]. Similarly, bvFTD patients had higher volumes than aAD patients in posterior [t(69.6)=2.45, p=0.029] but not anterior hippocampus [t(57.7)=-1.51, p=0.16].

## DISCUSSION

This study characterizes bvAD patients with known or likely AD pathology by comparing their neuropsychological profile and distribution of neocortical and hippocampal atrophy to bvFTD patients, aAD patients, and elderly controls. Patients with bvAD exhibited a blend of features of both bvFTD and aAD, and were significantly worse than controls on all measures. Results confirmed executive impairments in bvAD, showing that bvAD were as impaired on executive function measures as bvFTD patients and more impaired than aAD. They also displayed mild-to-moderate visuospatial processing and memory impairments; bvAD patients were significantly more impaired on visual copy and recall tasks than bvFTD patients, but not as impaired as aAD patients. bvAD patients exhibited significantly higher rates of agitation than bvFTD patients and of disinhibition and motor disturbance than aAD. MRI comparisons revealed that bvAD patients had cortical thinning relative to bvFTD patients centered in temporal-occipital regions, and cortical thinning relative to aAD patients mostly in frontal-temporal regions with a bias towards right hemisphere disease. bvFTD patients had cortical thinning relative to bvAD patients in inferior frontal cortex. While both bvAD and bvFTD patients demonstrated hippocampal atrophy relative to controls, posterior hippocampal volume loss was less severe than in aAD for both groups. Furthermore, bvFTD patients had greater anterior hippocampal atrophy than bvAD patients.

In our comparative study, we found a neuropsychological impairment profile in bvAD that includes characteristics of both bvFTD and aAD. We found evidence of executive dysfunction in bvAD equivalent to that of bvFTD, reasonably consistent with other studies. In one study, patients with bvAD performed worse than bvFTD on composite scores of executive function [9], while two groups found no differences between bvFTD and a behavioral [28] or dysexecutive [26] AD group. Another study found that executive performance depended on the task: bvAD patients performed worse than (Trails, Stroop inhibition, design fluency), better than (repetitions in design fluency task), or the same as bvFTD patients (D-words, rule violations in design fluency task) [27]. Our results align with a recent meta-analytic review [57] that found that bvAD executive function was comparable to bvFTD and worse than aAD. Similar to some but not all previous studies, we also found that bvAD had visuospatial function and memory worse than bvFTD [26–28], but better than aAD. While two studies found comparable figure copy and recall performance between behavioral variant groups [26,28], two found worse visual memory in bvAD [9,27]. When comparing language impairment and verbal memory, we found no differences between bvAD and bvFTD or aAD. Three past studies also found no differences between bvAD and bvFTD in neuropsychological tests of verbal memory [26–28], although one of these described more severe language impairments in clinical assessments for bvAD [27]. Another study reported worse verbal memory in bvAD [9]. Our findings are relatively consistent with the recent meta-analysis [57] that combined verbal and visual memory tests and found that bvAD had a memory profile similar to aAD and marginally but not significantly worse than bvFTD.

The basis for a performance profile showing some visual memory impairment but less verbal memory deficits is unclear. One possibility is that right hemisphere disease tends to be associated more strongly with behavioral changes as well as visuospatial deficits, and as can be seen in Figure 3, bvAD patients tended to have greater right hemisphere disease than the other patient groups. Nevertheless, it should be kept in mind that conclusions differ for whether visuospatial deficits are a feature of bvAD; the current study found that bvAD have significantly worse performance on the Rey Figure copy and recall than bvFTD, while others report no difference [26–28]. Discrepancies in findings could be due in part to methodological differences, including using the less complex Benson Figure [27,28], a shorter delay (3 vs. 10-15 minutes) [26], or inclusion of patients with co-occurring TBI [9]. Additional work is needed to address this issue, since the relative deficit in visuospatial functioning seems to be an important distinguishing feature in comparisons of bvAD and bvFTD.

Our results showed selectively more frequent psychiatric and social impairment in bvAD. One possible speculation is that greater agitation in bvAD may be related in part to the visuospatial deficit in these patients, where challenges associated with interpreting the visual environment may provoke agitation in bvAD from a unique source that is not apparent in bvFTD. While the observation of greater neuropsychiatric impairment in bvAD is consistent with some studies, findings have been quite varied. In bvAD, more delusions and agitation [28,57,58], hallucinations [57,58], irritability, sensitivity, and emotional lability [12], or depression [27] have been observed, while other researchers found that bvFTD showed more agitation, anxiety and irritability [9] or apathy, aggression, and lack of insight [27]. Further work is needed to assess the neuropsychiatric profiles of bvAD and bvFTD patients and other instruments may offer more sensitivity to differences than the NPI.

We measured cortical thickness and hippocampal volume comparatively in bvAD, bvFTD, aAD, and healthy controls. Consistent with prior findings [15], we found that bvAD had more occipital, posterior temporal, and parietal lobe atrophy than bvFTD; and that bvFTD had more frontal and anterior temporal lobe atrophy than bvAD [9,26,27]. We also found that bvAD had more atrophy in prefrontal cortex and temporal cortex, with a bias towards right hemisphere disease, compared to aAD, supported by previous findings [9,26]. Using a novel algorithm for prior-based segmentation of medial temporal lobe structures from T1-weighted MR images, we found that bvAD and bvFTD patients exhibited posterior hippocampal atrophy that was intermediate between controls and aAD. In anterior hippocampus, bvFTD patients had significantly smaller volumes than bvAD patients and did not differ from the aAD group.

Our neuroimaging findings fit with the overall patterns of atrophy that have emerged from past research. Investigators in one study found that, regardless of neuropathological diagnosis, patients with a possible clinical diagnosis of bvFTD shared atrophy in the anterior cingulate, frontal insula, striatum, and amygdala [27]. This suggests that the degeneration of these regions is closely associated with the behavioral syndrome. It is noteworthy that one area of inferior frontal disease revealed cortical thinning in bvFTD relative to bvAD, and this may have contributed in part to the subtle behavioral differences between these two groups. More importantly, our study revealed areas of significant temporal-occipital atrophy in bvAD compared to bvFTD. While other studies have shown greater precuneus atrophy in bvAD than in bvFTD, we did not find this on direct comparison. However, our aAD group had lower cortical thickness than bvFTD, but not bvAD, in the precuneus and posterior cingulate gyrus. This is consistent with studies identifying the precuneus [59] and the posterior cingulate gyrus [60] as hubs of disease in patients with underlying AD pathology. One study limited its analysis to prefrontal and medial temporal regions, and found prefrontal and medial temporal lobe atrophy in both bvAD and bvFTD [26]. When directly comparing the two, they found no areas where bvAD had more atrophy than bvFTD and found that bvFTD patients had more frontal and temporal atrophy than bvAD. Comparing bvAD and aAD, their results matched ours: they found no areas where aAD had greater atrophy and found more frontal lobe atrophy in bvAD [26]. Researchers in another group observed that bvAD showed a predominantly temporoparietal pattern of atrophy similar to aAD, with additional deterioration in the left orbitofrontal cortex, frontal poles and middle and superior frontal gyri [9]. They also found that bvAD showed less frontal lobe involvement than bvFTD and suggested that bvAD has a more posterior distribution of atrophy than bvFTD [9]. We find this to be generally true in a direct comparison of bvAD and bvFTD groups. However, relative to aAD, both bvAD and bvFTD patients have more severe frontal atrophy; and relative to healthy controls, both groups have extensive atrophy throughout all lobes of the brain, with some sparing of early visual and sensorimotor cortex. Recently, authors reviewed the bvAD neuroimaging literature and suggested two distinct bvAD neuroanatomical phenotypes: one with more anterior atrophy (“bvFTD-like”) and one with relative frontal sparing (“AD-like”), with the latter being more prevalent [57].

Taken together with the results of previous studies, the current study lends support to the possible role of degeneration in posterior cortical regions as an anatomical marker to distinguish bvAD and bvFTD. This would also be consistent with the observation of relatively greater visuospatial deficits in bvAD than bvFTD. Furthermore, more anterior hippocampal atrophy in bvFTD and relatively greater posterior hippocampal atrophy in bvAD could also aid in differentiating bvAD from bvFTD, although relatively small samples prevented us from conclusively testing this hypothesis. Additional research comparing these groups’ hippocampal volumes will help clarify this finding’s potential significance.

This study had a number of limitations. First, bvAD is a relatively rare syndrome and we selected cases based on strict criteria for known or likely pathology. Thus, while our bvAD cohort was comparable to that of other studies, its small size likely affected our statistical power to detect neuropsychological (n ranging from 13 to 15 depending on test) and neuroanatomical (n=12) differences between bvAD and bvFTD. This is a challenge that previous studies have faced as well, since only a fraction of patients with a clinical diagnosis of bvFTD ultimately have AD pathology [18,19]. Second, only a subset of our patients had autopsy-confirmed pathology, and it is possible that some bvAD patients who were positive for AD using CSF biomarkers could have had both AD and FTLD pathologies since CSF markers to detect secondary FTLD pathology are still lacking. Third, in this exploratory study we compared bvAD to bvFTD and aAD across a battery of neuropsychological evaluations but did not adjust statistical analyses for comparisons across multiple tasks. While this increases the possibility of type I error, it is crucial to be able to identify differences across rare patient groups with similar phenotypes, and we are careful to interpret our results in the context of previous findings. Fourth, since our data were collected over many years, different subsets of subjects completed each neuropsychological task. Fifth, we note that we did not assess subcortical regions. Lastly, our samples consisted of mostly non-Hispanic White participants, which limits the generalizability of results for diverse racial and ethnic populations. Previous studies have found that non-amnestic [61] and dysexecutive [62] phenotypes, though rare, may be more frequent in African Americans than White Americans. Future work is needed to further clarify these potential differences and test if and how race and ethnicity interact with clinical presentation within the AD spectrum.

These limitations, along with other factors, may have contributed to a final caveat of our findings: there were no cognitive or behavioral assessments that showed greater impairment in bvFTD than bvAD. Although groups were matched on disease duration, FTLD is associated with faster progression than AD [63]. Therefore, those with FTLD pathology were presumably more advanced in their disease course relative to those with AD pathology. Patients with bvFTD, however, performed better on our measure of disease severity (MMSE). While difficult to assess, differences in disease severity may have influenced findings. Another potentially impactful variable is age. Our bvAD group was older at disease onset (median=63.5 years) than our bvFTD group (median=59 years), a pattern consistent with previous findings [26–28]. While the age at onset in bvFTD is usually between 50 and 65 [64–67], studies have reported average onset ages ranging from 63 to 72 in bvAD [9,10,28]. To account for these possible influences, our analyses covaried for MMSE and age at test and, still, we did not see worse neuropsychological performance in bvFTD compared to bvAD. This is unexpected, given our neuroimaging results, which show decreased anterior hippocampal and inferior frontal volumes in bvFTD compared to bvAD. Perhaps more neuropsychological measures or a larger sample size would reveal relative impairments in bvFTD. Moreover, longitudinal studies may be best to understand the role of age of onset and disease duration and severity in clinical presentations and to characterize the core features and disease progression of bvAD. It is also important to consider that, when studying patients with a behavioral syndrome, poor test performance cannot always be attributed to cognitive deficits, but instead may reflect behavioral symptoms and diminished cooperation with testing procedures [68].

In summary, our study sought to profile an understudied variant of AD, bvAD, and to improve its discrimination from bvFTD and aAD by outlining the differences in behavioral, cognitive, and neuroanatomical features compared to these syndromes. We found evidence that supports bvAD as a somewhat distinct clinical syndrome, featuring identifiable selection characteristics seen in both bvFTD and aAD. Results revealed that the bvAD group showed more impairment on visuospatial copy and recall and had more agitation than the bvFTD group, and more behavioral and executive impairments with less visuospatial difficulty than aAD. The bvAD group had more posterior temporal-occipital atrophy, while bvFTD patients had more frontal and anterior temporal atrophy. bvAD patients also had more frontotemporal atrophy than aAD. Additionally, there was less anterior hippocampal atrophy in bvAD than bvFTD. Improving our understanding of the associations between pathology and clinical features in these groups will have critical implications for diagnosis, prognosis, caregiver burden, and effective treatment.

## Supporting information

Dominguez Perez STROBE checklist

## Data Availability

All data produced in the present study are available upon reasonable request to the authors.

## ACKNOWLEDGEMENTS

The authors would like to give special thanks to the research participants and research specialists at the Penn FTDC and Alzheimer’s Disease Center, who played an instrumental role by contributing to the data collected and analyzed in this study.

